# Estimations of SARS-CoV-2 endemic characteristics

**DOI:** 10.1101/2023.01.24.23284980

**Authors:** Igor Nesteruk

## Abstract

The fourth year of the COVID-19 pandemic without decreasing trends in the global numbers of new daily cases, high numbers of circulating SARS-CoV-2 variants and re-infections together with pessimistic predictions for the Omicron wave duration force studies about the endemic stage of the disease. The global trends were illustrated with the use the accumulated numbers of laboratory-confirmed COVID-19 cases and deaths, the percentages of fully vaccinated people and boosters and the results of calculation of the effective reproduction number provided by Johns Hopkins University. The modified SIR model showed the presence of unsteady equilibrium. The global numbers of new daily cases will range between 300 thousand and one million, daily deaths – between one and 3.3 thousand.

---

The fourth year of the COVID-19 pandemic, the high numbers of circulating SARS-CoV-2 variants [1-3] and re-infected persons [4-6], the lack of decreasing trends in the global numbers of new daily cases [7, 8], and the expected very long duration of the Omicron wave [9] make us think about the constant circulation of the pathogen, that is, about the endemic stage of the disease. To illustrate the global trends, let us use the accumulated numbers of laboratory-confirmed COVID-19 cases *V*_*j*_ and deaths *D*_*j*_, the percentages of fully vaccinated people *VC*_*j*_ and boosters *BC*_*j*,_ and the results of calculation of the effective reproduction number *R*_*j*_ [10]. Information corresponding to day *t*_*j*_ (starting with January 22, 2020) is available in COVID-19 Data Repository by the Center for Systems Science and Engineering (CSSE) at Johns Hopkins University (JHU) [8].

We will use the version the JHU file updated on December 7, 2022 and the smoothed accumulated characteristics, [11]:

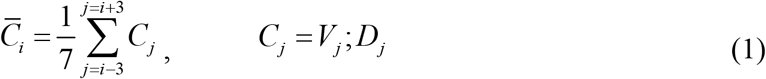

To estimate the smoothed numbers of new daily cases *DV*_*i*_, deaths *DD*_*i*_, and the case fatality risk *CFR*_*i*_ *=DD*_*i*_ */DV*_*i*_, the numerical derivatives of the smoothed values (1) are calculated as follows, [11]:

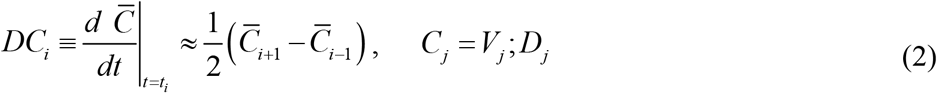

The solid lines in Fig.1 illustrate the vaccination levels *VC*_*j*_ and *BC*_*j*_ (green and yellow colors, respectively), *R*_*j*_ values (magenta) and calculations of *DV*_*i*_, *DD*_*i*_ and *CFR*_*i*_ (blue, black and red, respectively). It can be seen that after October 2020, smoothed global numbers of new daily cases were never less than 300,000. After April 2020, the effective reproduction number [10] was close to the critical value 1.0 [12-15]. In 2022 we see the sufficient drop in *CFI* figures. To estimate the long term trends we calculated average values of *DV, DD, CFI* for every year of the pandemic with the use of accumulated characteristics on January 22 and December 31, 2020; January 1 and December 31, 2021; and January 1 and December 6, 2022. The results are listed in Table 1 and shown in Fig. 1 by dashed lines.

**Table 1.** Averaged characteristics of the COVID-19 pandemic in 2020, 2021 and 2022.

| Year | Average daily numbers of new cases, $DV$ | Average daily numbers of deaths, $DD$ | Average case fatality risk, $CFD$ |
| --- | --- | --- | --- |
| 2020 | $2.4352e+005 = (83773024-557)/344$ | $5.5281e+003 = (1901671-17)/344$ | $0.0227 = (1901671-17)/(83773024-557)$ |
| 2021 | $5.6155e+005 = (288738770-83773024)/365$ | $9.7745e+003 = (5469351-1901671)/365$ | $0.0174 = (5469351-1901671)/(288738770-83773024)$ |
| 2022 | $1.0516e+006 = (646279687-288738770)/340$ | $3.4567e+003 = (6644643-5469351)/340$ | $0.0033 = (6644643-5469351)/(646279687-288738770)$ |

**Fig. 1.**
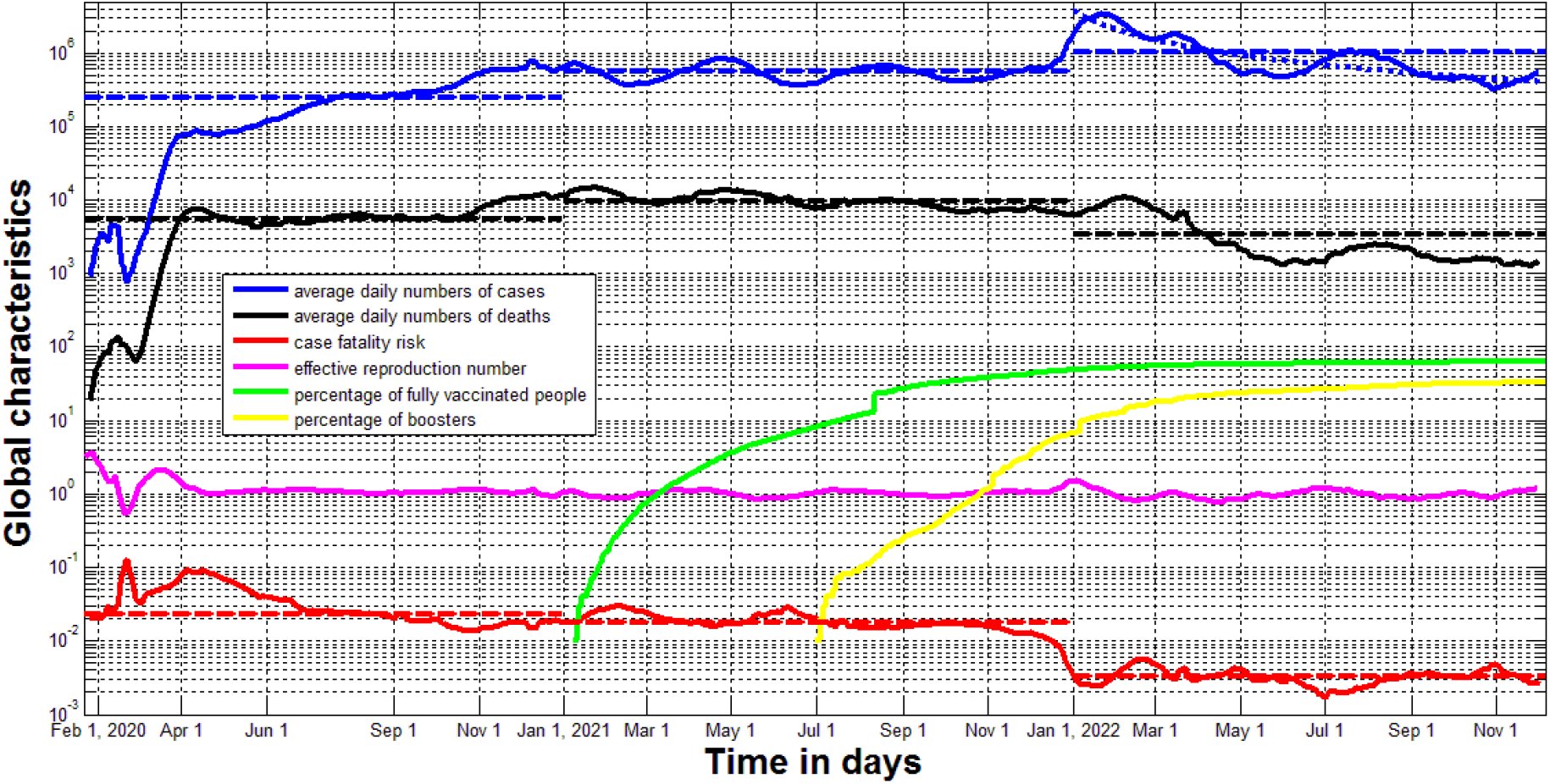
Global characteristics of the COVID-19 pandemic. Green, yellow and magenta lines respectively show *VC*_*j*_, *BC*_*j*_, *R*_*j*_ values listed in [8]. Solid blue, black and red represent calculations of *DV*_*i*_, *DD*_*i*_ and *CFR*_*i*_, respectively (eq. (1) and (2)). Corresponding dashed lines represent the average values for different years listed in Table 1. The dotted blue line shows the results of non-linear correlation (eq. (9)).

The *DV* values approximately doubled every year, the *DD* figure in 2021 was 1.8 times higher than in 2020. But in 2022 we see sufficient decrease in mortality and much lower *CFR* values. This can be explained as a positive effect of vaccinations (the global VC values tend to some saturation in 2022, see green line), lower pathogenicity of the Omicron strain (which began to spread widely at the end of 2021, [16, 17]), as well as the influence of natural immunity [18].

The classical SIR-model [19-24] relates the numbers of susceptible *S(t)*, infectious *I(t)* and removed persons *R(t)* over time *t* and can be successfully used for simulations of the first COVID-19 waves in individual countries and worldwide [11, 25-27]. Numerous improvements of this model [9, 11, 18, 28, 29] do not take into account re-infections, which can be simulated by adding terms ±*δ R* to the first and last differential equations:

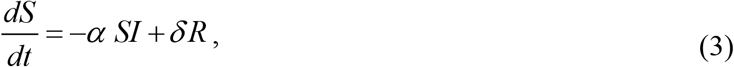

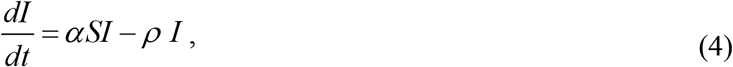

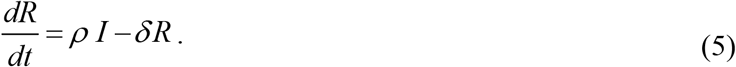

Now compartment *S(t)*, which includes the persons who are sensitive to the pathogen, is increasing with rate *δ R* due to the persons who have lost their immunity and can be reinfected. The same term with opposite sign appears in eq. (5).

The set of differential equations (3)-(5) has the equilibrium point [30-32] (*S*_*\**_,*I*_*\**_,*R*_*\**_) corresponding to zero values of the derivatives:

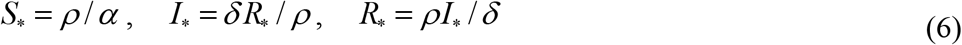

At the equilibrium, the daily number of new cases *DV*_*\**_ must be equal to the daily number of new infections; to the sum of immunized, isolated and dead persons; and to the number of persons who have lost their immunity and move from compartment *R* to *S*, i.e.:

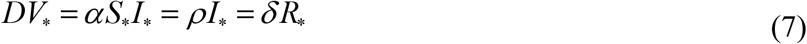

The Jacobian matrix [30-32] of set (3)-(5) can be written as follows:

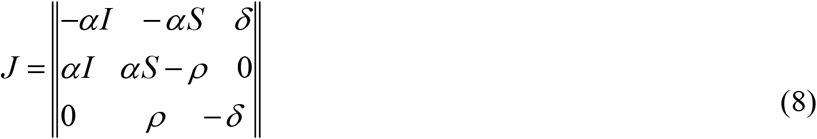

Taking into account (6), the eigenvalues of (8) at the equilibrium point can be calculated as follows:

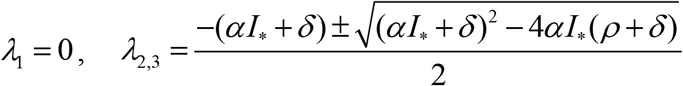

The presence of a zero eigenvalue indicates that the equilibrium cannot be asymptotically stable, that is, the system does not approach equilibrium with increasing time and we will always observe some oscillations.

To estimate the endemic global number of new daily cases we can use the average *DV* value for 2022 (around 1 million), presented in Table 1. More optimistic prediction *DV* ≈ 300, 000 follows from the minimum of *DV*_*i*_ values illustrated by the blue solid line and non-linear correlation approach [33], which yields the best fitting curve

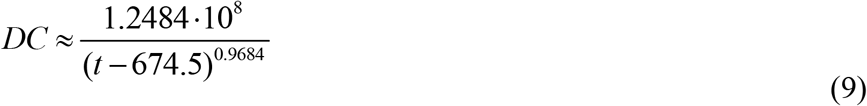

for the 2022 dataset (shown by the dotted blue line). Thus, the global daily numbers of SARS-CoV-2 cases can vary from 300 thousand to one million and the daily numbers of deaths from 1.0 to 3.3 thousand (if we use the *CFR* value from the Table 1 corresponding to 2022). According to the recent WHO report “nearly 2.8 million new cases and over 13 000 deaths were reported in the week of 9 to 15 January 2023. In the last 28 days (19 December 2022 to 15 January 2023), nearly 13 million cases and almost 53 000 new deaths were reported globally”, [7]. These figures are consistent with the given estimates of SARS-CoV-2 endemic characteristics.

It is necessary to emphasize that the presented estimates refer to the laboratory confirmed cases only. Due to the high numbers of asymptomatic patients and insufficient testing level, the real number of SARS-CoV-2 cases is much higher [33-35]. To estimate the global visibility coefficient let us take the number of cases per million 72,524 registered worldwide as of August 1, 2022 [8]. On the same day, the average value was 460,834 for European countries with high enough testing level (more than 3 tests per capita) [33]. It means that the real numbers of cases can be approximately 6 times higher than registered figures. In the future, reducing the testing level can lead to lower the numbers of detected cases and deaths.

Other restrictions on the accuracy of the given estimates may be related to the situation in mainland China, where the Zero-COVID policy [36] began to be lifted only after November 30, 2022 [37]. Very high numbers of new daily cases in December 2022 reported by media [38] does not look reliable [39], nevertheless it is better to use the presented estimations excluding the Chinese datasets.

According to the presented estimations, the annual mortality caused by SARS-CoV-2 will range from 365 thousand to 1.2 million. Unfortunately, these figures are higher than in the case of seasonal influenza (between 294 and 518 thousand annual deaths in the period from 2002 to 2011, [40]).

## Data Availability

All data produced in the present work are contained in the manuscript

## Acknowledgement

The author is grateful to Tetsuro Kobayashi and Oleksii Rodionov for their help in providing very useful information.

